# Bronchoscopy in critically ill COVID-19 Patients: microbiological profile and factors related to nosocomial respiratory infection

**DOI:** 10.1101/2020.07.01.20144683

**Authors:** Pere Serra, Carmen Centeno, Ignasi Garcia-Olivé, Adrià Antuori, Maria Casadellà, Rachid Tazi, Fernando Armestar, Ester Fernández, Felipe Andreo, Antoni Rosell

## Abstract

**Background:** Nosocomial co-infections are a cause of morbidity and mortality in Intensive Care Units (ICU).

**Objectives:** Our aim was to describe bronchoscopy findings and analyse co-infection through bronchial aspirate (BA) samples in patients with COVID-19 pneumonia requiring ICU admission.

**Methods:** We conducted a retrospective observational study, analysing the BA samples collected from intubated patients with COVID-19 to diagnose nosocomial respiratory infection.

**Results:** One-hundred and fifty-five consecutive BA samples were collected from 75 patients. Of them, 90 (58%) were positive cultures for different microorganisms, 11 (7.1%) were polymicrobial, and 37 (23.7%) contained resistant microorganisms. There was a statistically significant association between increased days of orotracheal intubation (OTI) and positive BA (18.9 days versus 10.9 days, p<0.01), polymicrobial infection (22.11 versus 13.54, p<0.01) and isolation of resistant microorganisms (18.88 versus 10.94, p<0.01). In 88% of the cases a change in antibiotic treatment was made.

**Conclusion:** Nosocomial respiratory infection in intubated COVID-19 patients seems to be higher than in non-epidemic periods. The longer the intubation period, the greater the probability of co-infection, isolation of resistant microorganisms and polymicrobial infection. Microbiological sampling through BA is an essential tool to manage these patients appropriately.

## INTRODUCTION

Coronavirus disease 2019 (COVID-19) is an unprecedented outbreak of potentially severe pneumonia^1^, and about 5% of patients with COVID-19 require management in an intensive care unit (ICU)^2,3^. These patients are at a high risk of developing multiple types of secondary pulmonary infection and therefore a bronchial culture would be needed to elucidate the causal agent^4,5^.

Bronchoscopy is a widely used technique in critically ill patients^6^. Although the evidence of its role during the COVID-19 pandemic is sparse and the risk of infection for health-care workers is high^7,8^, it has been shown to be necessary to manage complications, as well as to obtain samples for microbiological cultures and the sensitive detection of SARS-CoV-2^9,10^. It is also essential to assist in the management of artificial airway procedures^9^. Furthermore, although the diagnosis of ventilator-associated pneumonia remains notoriously difficult, it occurs in up to ∼30% patients (20–60% of suspected cases)^11^, and accurate sampling can reduce the unnecessary use of antibiotics^3^.

The aim of this study is to describe the bronchoscope findings, the microbiological profile and its characteristics, and the factors related to nosocomial respiratory co-infections in critically ill COVID-19 patients.

## METHODS

The study is retrospective and observational, analysing the BA samples collected from intubated patients with COVID-19 to diagnose nosocomial respiratory infection. It was conducted at the referral Hospital Germans Trias i Pujol in Badalona (Barcelona, Spain), which covers a population of over 800,000 inhabitants with 600 beds, including 52 critically ill beds, which were expanded to 150 during the COVID-19 pandemic.

### Study subjects

Patients hospitalized for severe infection with COVID-19 with mechanical ventilation in ICU who required a bronchoscopy and underwent a BA in order to confirm a nosocomial respiratory tract infection were included.

### Bronchoscopy performance

Procedures were performed with disposable scopes (Ambu⍰ aScope4™ Broncho, Large OD 5.8 mm/ID 2.8 mm or Regular OD 5.0 mm/ID 2.2 mm, Copenhagen, Denmark) in patients under usual intravenous sedation with muscle relaxant, through orotracheal tube or tracheostomy, in pressured controlled ventilation mode. Only the minimal staff required attended the procedure and all personnel wore personal protective equipment, FFP3 masks, double gloves and double eye protection and appropriate hand washing was performed before and after the bronchoscopy^3,8,9^.

### Sample obtaining

BA was performed during the bronchoscope procedure by suctioning deep secretions with a new, sterile suction circuit. A minimum volume of 2-3ml of specimen was collected into a sterile, leak-proof container for microbiological sampling^7^. Gram stain and semi-quantitative cultures were performed for endotracheal aspirates. Isolated bacteria were identified by standard laboratory methods and susceptibility to antimicrobial agents was determined following European Committee on Antimicrobial Susceptibility Testing (EUCAST) procedures.

### Statistical analysis

Categorical variables are expressed as frequencies and interquartile range (IQR), while quantitative variables are expressed as the mean and standard deviation (SD). Association between variables of interest and binary outcomes was analysed using a logistic regression, while quantitative variables were compared using the t-student test.

We defined resistance as multidrug-resistant (MDR), extensively drug-resistant (XDR) and pandrug-resistant (PDR) bacteria, based on International standard definitions for acquired resistance^12^. For the analysis, we considered MDR, XDR and PDR as Resistant, and the rest were considered non-resistant. The Kaplan-Meier method was used to measure the fraction of patients with a positive culture of BA or Resistant microorganisms during their ICU stay. The association between the time of orotracheal intubation and microbiological results was assessed with a logistic regression. These analyses were conducted with version 15 of STATA.

### Ethical considerations

The research protocol was approved by the hospital’s COVID-19 committee (Reference 2342342/20).

## RESULTS

### Patients

Three-hundred and two successive bronchoscopies were performed on mechanically ventilated patients with COVID-19 in the ICU from 24 March to 15 May. Bronchoscopy was well tolerated and no complications occurred during the bronchoscopy procedure.

One-hundred and fifty-five successive BA samples were collected from 75 patients and were included in the analyses. Sixty-five (86.1%) of these patients were male and 10 (13.9%) were female, with a mean age of 59 years (SD 10.2). Only 10 (13.3%) of the patients had previous known respiratory pathologies, namely five cases of asthma and five cases of chronic obstructive pulmonary disease (COPD), and only one patient was immunosuppressed. Patient characteristics are shown in Table 1.

**Table 1.** Patient characteristics (n=75).

| Characteristics | Values |
| --- | --- |
| Male sex, n (%) | 65 (86.7) |
| Age, years, m | 59 ± 10.2 |
| Cardiovascular Risk, n (%) | 47 (62.6) |
| Respiratory pathologies, n (%) | 10 (13.3) |
| Obesity, n (%) | 42 (56) |
| Corticoids treatment, n (%) | 63 (84) |
| Tocilizumab treatment, n (%) | 37 (49.3) |
| Previous Antibiotic treatment, n (%) | 151 (96.8) |

The number of mean days between orotracheal intubation and the first bronchoscopy was 8.40, and the average number of bronchoscopies per patient was 2.04.

The predominant findings were the presence of abundant thick secretions, which were usually difficult to suction in 91% of the patients, and muco-haematic plugs normally in the tube or central airway, which required the use of saline and a mucolytic agent in 32% of the patients, especially in extracorporeal membrane oxygenation patients. After five days of intubation, we observed a mucous biofilm around the tracheal tube in the majority of patients.

### Microbiology

Ninety (58%) BA samples tested positive for different microorganisms, of which 11 (7.1%) were polymicrobial, rendering a total of 101 microorganisms detected. Thirty-eight (37.6%) contained resistant microorganisms: 30 (79%) were MDR, four (10.5%) were XDR and four (10.5%) were PDR. From all the positives cultures, 90 (89.1%) were bacterial and 11 (11.9%) were fungal (*Aspergillus spp or Candida albicans*). The microbiological results, adding the polymicrobials, were: 33 (19.7%) *Pseudomonas aeruginosa*, 17 (10.1%) *Enterobacter cloacae*, 9 (5.3%) *Klebsiella oxytoca*, 7 (4.1%) *Staphylococcus aureus*, 5 (2.9%) *Enterobacter spp*, 4 (2.3%) *Achromobacter xylosoxidans*, 4 (2.3%) *Klebsiella aerogenes*, 3 (1.7%) *Escherichia coli*, 2 (1.1%) *Citrobacter spp*, 2 (1.1%) *Klebsiella pneumoniae*, 3 (1.7%) *Morganella morganii*, 1 (0.5%) *Serratia marcescens*, and 8 (4.7%) *Candida albicans* and 3 (1.7%) *Aspergillus spp*.

Eighty (88.8%) of the positive cultures involved a change or a new treatment, while only 10 (11.2%) of them did not, for different reasons. Three subjects had *Candida albicans* and two subjects had sensitive *Staphylococcus aureus*, which were all considered to be non-clinically significant. Of the remaining patients, two had *Pseudomonas aeruginosa*, one had *Enterobacer cloacae* and two had *Klebsiella pneumoniae*, and they were already in treatment following a previous positive BA sample. In three cases with previous negative nasopharyngeal swabs tests, the BA sample tested positive for COVID-19.

### Factors related to nosocomial infection

We found a statistically significant association between days of orotracheal intubation (OTI) and positive BA (18.9 days versus 10.9 days, p<0.01), polymicrobial infection (22.11 versus 13.54, p<0.01) and isolation of resistant microorganisms (18.88 versus 10.94, p<0.01). The odds ratios (OR) for the associations can be seen in Table 2.

**Table 2.** Results of logistic regressions describing the association between days of intubation and microbiological findings.

|  | Odds ratio | 95% CI | p-value |
| --- | --- | --- | --- |
| Positive | 1.07 | 1.03-1.11 | p=0.000 |
| Polymicrobial | 1.05 | 1.01-1.09 | p=0.009 |
| Resistant | 1.05 | 1.02-1.08 | p=0.001 |

The risk of having a positive culture at day 10, 15, 20 and 25 after OTI was 24.5 (95% CI 17.0-34.7), 36.4 (95% CI 26.6-48.5), 62.1 (95% CI 49.2-75.1) and 76.2% (95% CI 62.6-87.6), respectively (Figure 2). The risk of isolation of a MR microorganism at day 10, 15, 20 and 25 after OTI was 4.8 (95% CI 2.0-11.2), 12.3 (95% CI 6.7-22.1), 22.6 (95% CI 13.8-35.8) and 30.5% (95% CI 18.7-47.2), respectively (Figure 3).

**Figure 1.**
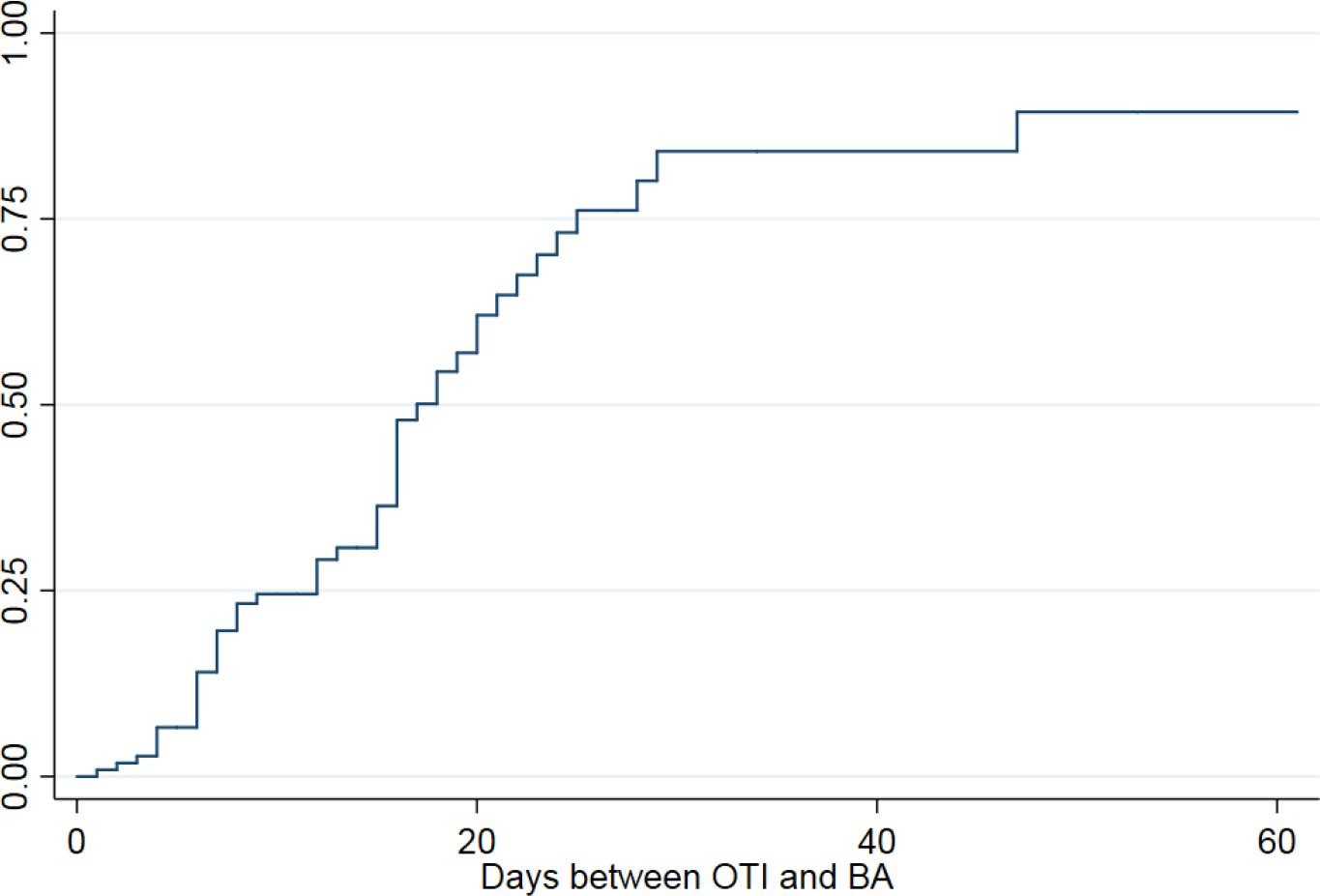
Kaplan-Meier curve showing the probability of presenting a positive-culture BA according to the days since OTI.

**Figure 2.**
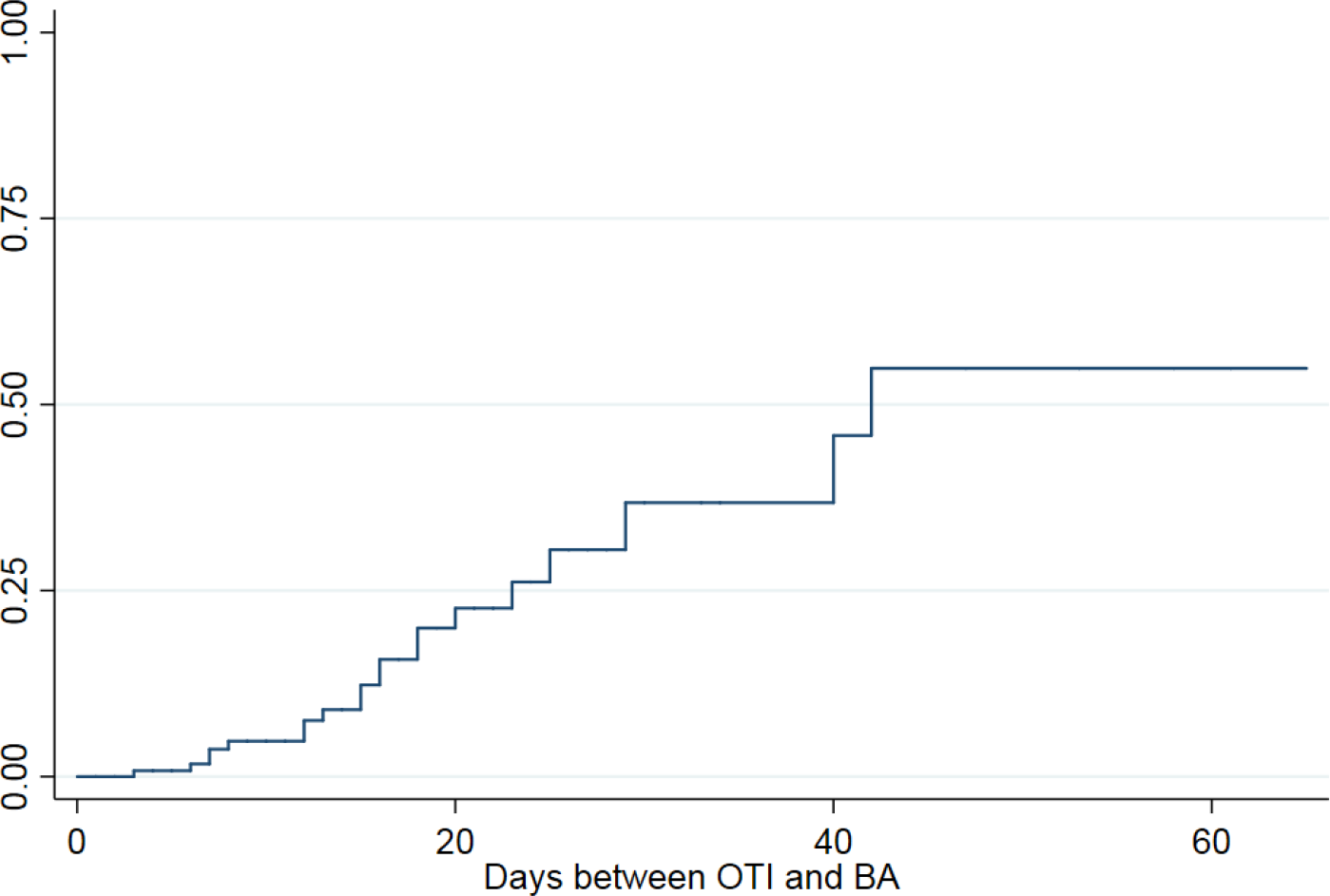
Kaplan-Meier curve showing the probability of presenting a positive-MR-isolation in BA samples according to the days since OTI.

Neither corticoids, tocilizumab treatment nor previous antibiotic treatment were associated with an increased risk of positive BAS, polymicrobial result or resistant microorganisms, as analysed with a logistic regression. Neither were having a previous respiratory pathology or a cardiovascular risk factor were not associated with an increased risk either.

## DISCUSSION

The most frequent bronchoscopy findings in patients with severe COVID-19 in the ICU were the presence of abundant thick secretions, usually difficult to suction, requiring more saline and mucolytic agents than usual and hematic plugs. This may be due to the use of heat and moisture exchange filters close to the patient instead of a heated humidified circuit^13^ or the average ICU stay, but as an author suggested^9^, we cannot ascertain whether thick secretions can be attributed to the virus infection itself.

Our data show that more than a half of the BA samples obtained by bronchoscopy in critically ill COVID-19 patients were positive. The incidence of co-infection (58%) seems to be higher than usual in non-COVID-19 epidemic periods (∼ 30%)^14,15^, as suggested by Kim et all^16^ in a study on nasopharyngeal swabs of symptomatic patients. To the best of our knowledge, to date there is only one published letter to the editor about bronchoscopies in critically ill COVID-19 patients, with 63 mini-BAL, in which they only found 28.6% of co-infection^9^. The average ICU stay in our series is longer, which may explain the number of co-infections. The morbidity and mortality rates remain high in co-infection and the microbial aetiology is variable, so the etiological diagnosis may be of vital importance^15,17^.

*Pseudomonas aeruginosa* is the most frequent microorganism, according to the latest releases of ventilator-associated pneumonia^15^, but the microbiological isolations had some differences with samples obtained in non-epidemic periods^9,14,17^. Zhou et al^18^ showed that in the current COVID-19 pandemic, 50% of infected patients who died had secondary bacterial infections, but no information is available regarding the antimicrobial sensitivities of the organisms that were identified^18^.

Our study also provides data regarding antimicrobial sensitivities and resistances, which could provide new knowledge that may help to change clinical management. And indeed, in our experience, this involved a change or a new treatment in almost 90% of patients with positive cultures. As we have previously commented, accurate sampling can reduce the unnecessary use of antibiotics^3^. By assessing the risk, bronchoscopy can help to improve these patients and will improve antimicrobial stewardship throughout the course of the pandemic^4,7^.

Furthermore, the longer the intubation period, the higher the probability of co-infection and of developing resistant microorganisms, which were both statistically significant associations. From the tenth day of IOT, the risk of co-infection increases steadily, and these can lead to increased disease severity and mortality^4^.

Neither corticoids, tocilizumab treatment nor previous antibiotic were associated with an increased risk of positive BA, polymicrobial result or resistant microorganisms. Despite the frequent prescription of broad-spectrum empirical antimicrobial drugs in patients with coronavirus associated respiratory infections, there is no evidence to support its association with respiratory co-infection^19^. Future prospective evidence is needed to support these data.

In conclusion, co-infection, diagnosed with BA, in intubated COVID-19 patients seems to be higher than in non-epidemic COVID-19 periods. The probability of co-infection and of developing resistant microorganisms and polymicrobial infections becomes significantly higher at the tenth day after the OTI. Bronchoscopy microbiological sampling is an important tool in co-infection diagnosis and the clinical management of these patients and also for co-infection diagnosis.

## Data Availability

The data is Availability for any revision.

## Statement of ethics

The manuscript has not been published or presented elsewhere in part or in entirety, and it is not under consideration by another journal. All study participants provided informed consent, and the study design was approved by the appropriate ethics review board. We have read and understood your journal’s policies, and we believe that neither the manuscript nor the study violates any of these.

## Conflicts of Interest Statement

The authors have no conflicts of interest to declare.

## Funding sources

The authors have no funding sources to declare Disclosure Statement.

## Author contribution

Study design: PSM, CCC.

Bronchoscopy performance: PSM, CCC, RTM, EFA, FAG.

Data collecting: PSM, CCC.

Statistical analysis: IGO, PSM

Wrote the manuscript: PSM, CCC, IGO, MCF.

Read critically the manuscript: FAR, AAT, FAG, ARG.

## Notes

### Competing Interest Statement

The authors have declared no competing interest.

### Funding Statement

There are no funding supported in the work presented

### Author Declarations

The research protocol was approved by the hospital's COVID-19 committee (Reference 2342342/20)

